## Supplementary File for "Large-scale seroepidemiology uncovers nephro-urological pathologies in persons with tau autoimmunity"

Supplemental Figure 1. Uncropped Western Blot images of Figure 2E

Supplemental Figure 2. Epitope mapping of patient samples shown in Figure 3.

Supplemental Figure 3. Risk ratio for tau autoantibodies in autoimmune diseases.

Supplemental Figure 4. Sequence alignment comparing the sequence of MTBD-tau (residues 13–114) to human NAMPT (residues 368–461).

Supplemental Table 1. Targeted AD screen samples.

Supplemental Table 2. Laboratory parameters used in the statistical analysis.

Supplemental Table 3. ICD-10 codes used for the grouping of neurological disorders in the statistical analysis.

Supplemental Table 4. ICD-10 codes used for the grouping of systemic disorders in the statistical analysis.

**Supplemental Figures**

**Supplemental Figure 1. Uncropped Western Blot images of Figure 2E.** Unmodified Western blot images of loading control (Actin) with molecular weight standard. As positive control the anti-tau commercial RD4 antibody was used and as negative control the goat anti-Human IgG Fc-gamma specific antibody was used (IgG antibody only).

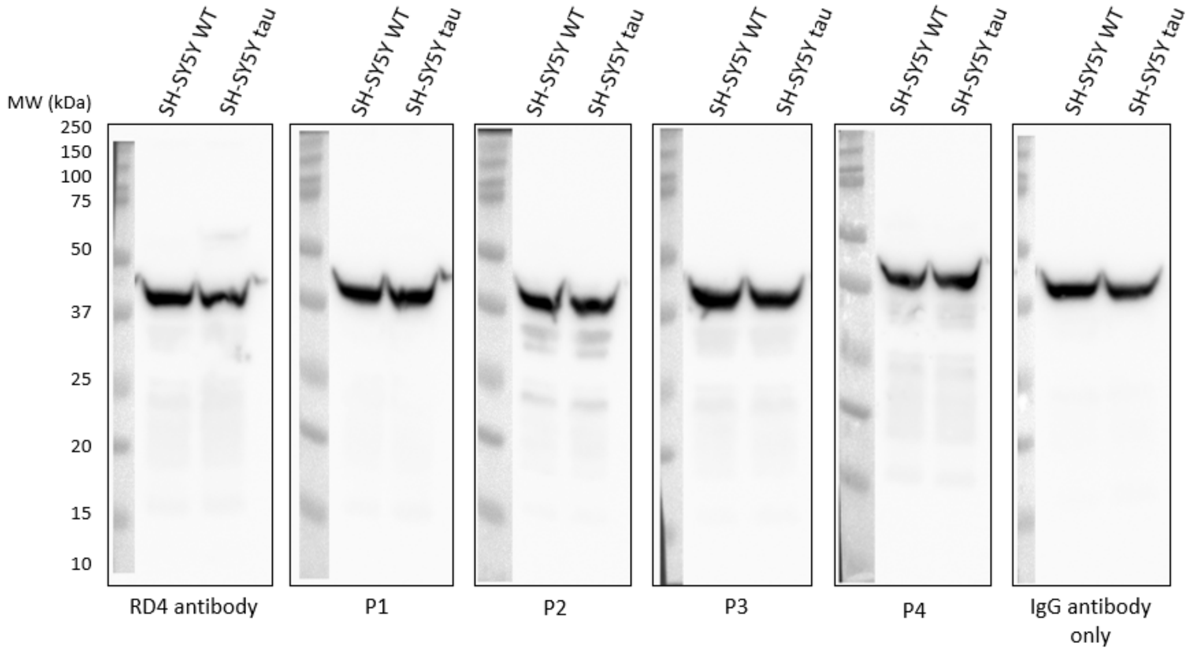

**Supplemental Figure 2. Epitope mapping of patient samples shown in Figure 3.** Epitope mapping of the same samples used in the assay shown in Figure 3 against eight 25mer MTBD-tau peptides overlapping 10 residues.

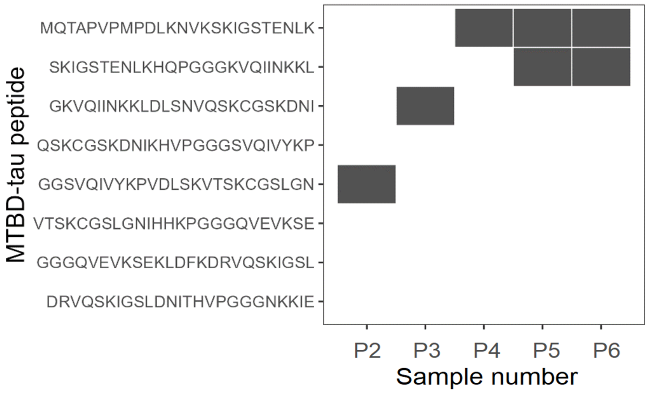

**Supplemental Figure 3. RR for tau autoantibodies in autoimmune diseases.** Forest plot showing the risk ratios and 95% CI (I bars) for the detection of tau autoantibodies in plasma samples of patients according to ICD-10 diagnosis of 5 different autoimmune diseases. aRR and 95% CI were estimated using log-binomial regression multivariate models including the respective variable, age and sex.

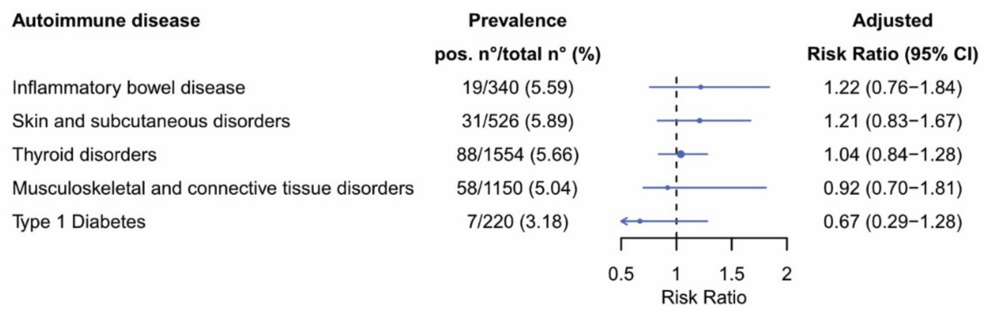

**Supplemental Figure 4. Sequence alignment comparing the sequence of MTBD-tau (residues 13–114) to human NAMPT (residues 368–461; P43490).** The alignment results from a BLASTP search with the BLOSUM62 substitution matrix (https://blast.ncbi.nlm.nih.gov/). The search was conducted against proteins presence in the human kidney and urinary proteome (UniProt/Swiss-Prot). The alignment revealed 28% sequence identity.

```
MTBD-tau  13   VKSKIGSTENLKHQPGGGKVQIINKKLDLSNVQSKCGSKDNIKHVPGGGSVQIVYKPVDL  72
               +K K+ S EN+    GGG +Q + +  DL N   KC        V  G  + +   PV
NAMPT     368  MKQKMWSIENIAFGSGGGLLQKLTR--DLLNCSFKCSYV-----VTNGLGINVFKDPVAD  420


MTBD-tau  73   SKVTSKCGSLGNIHHKPGGGQVEVKSEKLDFKDRVQSKIGSL  114
                   SK G L ++H  P G  V ++  K D ++  Q  + ++
NAMPT     421  PNKRSKKGRL-SLHRTPAGNFVTLEEGKGDLEEYGQDLLHTV  461
```

### Supplemental Tables

**Supplemental Table 1. Targeted AD screen samples.**

| | AD | Control | P |
|---|---|---|---|
| Women, n | 30 | 39 | |
| Men, n | 17 | 29 | |
| Age, median (IQR range) | 78 (70,5-86) | 81 (71-85) | 0.594 |

**Supplemental Table 2. Laboratory parameters used in the statistical analysis.**

| **Laboratory Parameter** | **Unit** |
|---|---|
| Sodium | mmol/L |
| Cholesterol, total | mmol/L |
| Chloride | mmol/L |
| C-reactive protein | mg/L |
| Iron | µmol/L |
| Cholesterol-LDL | mmol/L |
| Phosphate | mmol/L |
| Uric acid | µmol/L |
| Ferritin | µg/L |
| Glucose | mmol/L |
| Transferrin | µmol/L |
| Lipase | U/L |
| Myoglobin | µg/L |
| N-terminal pro-B-type natriuretic peptide (NT-pro-BNP) | ng/L |
| Parathyroid hormone | ng/L |
| Protein, urine | g/L |
| Protein-to-creatinine ratio, urine | g/mmol |
| Activated partial thromboplastin time (aPTT) | seconds |
| Erythrocytes | T/L |
| Mean corpuscular volume | fL |
| Mean corpuscular hemoglobin | pg |
| Reticulocytes | G/L |
| Hyperchromic erythrocytes | % |
| Hypochromic erythrocytes | % |
| Neutrophils | G/L |
| Monocytes | G/L |
| Eosinophils | G/L |
| Basophils | G/L |
| Lymphocytes | G/L |

| | |
|---|---|
| Large unstained cells | G/L |
| Leukocytes | G/L |
| Neutrophils | % |
| Monocytes | % |
| Eosinophils | % |
| Basophils | % |
| Lymphocytes | % |
| Large unstained cells | % |
| International normalized ratio (INR) | |
| Red cell distribution width (RDW) | % |
| Mean platelet volume | fL |
| Fibrinogen | g/L |
| Anti-factor Xa activity | IU/mL |
| IgG | g/L |
| IgA | g/L |
| IgM | g/L |
| Cholesterol-HDL | mmol/L |
| Lactate | mmol/L |
| Albumin-to-creatinine ratio, urine | mg/mmol |
| Potassium | mmol/L |
| Amylase | U/L |
| Lactate dehydrogenase | U/L |
| Albumin | g/L |
| Aspartate aminotransferase | U/L |
| Calcium, total | mmol/L |
| Urea | mmol/L |
| Magnesium | mmol/L |
| Protein | g/L |
| Creatinine | µmol/L |
| Free triiodothyronine (FT3) | pmol/L |
| Thyroid-stimulating hormone | mU/L |
| Glucose (fasting) | mmol/L |

| | |
|---|---|
| Anti-HBs | IE/L |
| Anti-HBc-IgG/IgM ratio | |
| Anti-HCV-IgG | |
| HIV Ag/Ab ratio | |
| Procalcitonin | µg/L |
| Bilirubin, total | µmol/L |
| Calcium, albumin corrected | mmol/L |
| HBs-Antigen | |
| pH, urine | |
| Density, urine | g/mL |
| Leukocytes, urine | /µL |
| Erythrocytes, urine | /µL |
| Renal tubular cells, urine | /µL |
| Squamous epithelial cells, urine | /µL |
| Hyaline casts, urine | /µL |
| Yeast, urine | /µL |
| Thrombin time | seconds |
| eGFR CKD-EPI 2009 | mL/min/1.73m$^2$ |
| Hemoglobin | g/L |
| Hematocrit | L/L |
| Mean corpuscular hemoglobin concentration | g/L |
| Hemoglobin distribution width (HDW) | g/L |
| Retikulozyten (automatisch) % | % |
| Immature granulocytes, absolute | G/L |
| Immature Granulocytes % (automatisch) | % |
| Nucleated red blood cells, absolute | G/L |
| Nucleated red blood cells | /100 Leukocytes |
| Transferrin saturation | % |
| Hemoglobin A1c, NGSP | % |
| Hemoglobin A1c, IFCC | mmol/mol |
| Gamma-glutamyl transferase | U/L |

| | |
|---|---|
| Alanine aminotransferase | U/L |
| Creatine kinase, total | U/L |
| Troponin T, high sensitivity | ng/L |
| eGFR BIS1 | mL/min/1.73m$^2$ |
| Prothrombin time | seconds |
| non-HDL-cholesterol | mmol/L |
| Triglycerides | mmol/L |
| Alkaline phosphatase | U/L |
| Free thyroxine (FT4) | pmol/L |
| 25-hydroxyvitamin D | µg/L |
| Osmolality | mmol/Kg |
| Vitamin B12 | ng/L |
| Ferritin (at risk indivuduals) | µg/L |
| Transferrin saturation (at risk individuals) | % |

**Supplemental Table 3. ICD-10 codes used for the grouping of neurological disorders in the statistical analysis.**

| Group of Disorders | ICD-10 codes |
|---|---|
| Alzheimer's disease | G30, F00 |
| Non-Alzheimer's dementia | F01, F02, F03, G31.0, G31.3, G31.82 |
| Epilepsy | G40, G41 |
| Migraine | G43 |
| Other headache disorders | G44, G50 |
| Meningitis, Encephalitis or Myelitis | G00, G01, G02, G03, G04, G05 |
| Parkinson's disease | G20 |
| Secondary parkinsonism | G21 |
| Atypical parkinsonism | G23.1, G23.2, G23.3, G23.8 |
| Essential tremor | G25.0 |
| Dystonia | G24 |
| Myoclonus | G25.3 |
| Multiple sclerosis | G35 |
| Neuropathies | G51, G52, G53, G56, G57, G60, G61, G62 |
| Myoneural junction disorders | G70 |
| Myopathies | G71, G72 |
| Hereditary ataxia/spastic paraplegia | G11 |
| Motor neuron disease | G12.2 |
| Huntington's disease | G10 |
| Sleep disorders | G47 |
| Stroke | G45 |
| Infarction | I63, G46, I69.3 |
| Hemorrhage | I60, I61, I62.9, I69.0, I69.2 |
| Alzheimer’s disease | G30.0, G30.1, G30.8, G30.9, F00.0, F00.1, F00.2, F00.9 |
| Frontotemporal dementia | G31.0 |
| Progressive supranuclear palsy | G23.1 |

**Supplemental Table 4. ICD-10 codes used for the grouping of systemic disorders in the statistical analysis**.

| Group of Disorders | ICD-10 codes |
|---|---|
| Cardiovascular | Ixx |
| Heart | I0x, I20, I21, I22, I23, I24, I25, I3x, I4x, I5x |
| Hypertension | I1x |
| Vascular | I7x |
| Digestive | Kxx |
| Stomach and intestine | K2x, K3x, K4x, K5x, K60, K61, K62, K63, K64 |
| Liver | K7x |
| Endocrine, Metabolic & Nutritional | Exx |
| Diabetes | E10, E11, E12, E13, E14 |
| Nutrition | E0x |
| Thyroid | E4x, E5x, E60, E61, E63, E64, E65, E66, E67, E68 |
| Hematopoietic | D5x, D6x, D7x, D8x |
| Anemia | D5x |
| Coagulation | D65, D66, D67, D68, D69 |
| Infections | Axx, Bxx |
| Kidney & Urinary | Nxx |
| Kidney | N0x, N1x, N2x |
| Urinary | N3x |
| Musculoskeletal | Mxx |
| Neoplasms | Cxx, D0x, D1x, D2x, D3x, D4x |
| Hematopoietic | C8x, C90, C91, C92, C93, C94, C95, C96 |
| Malignant solid | C0x, C1x, C2x, C3x, C4x, C5x, C6x, C71, C72, C73, C74, C75 |
| Neurologic | F0x, Gxx |
| Pregnancy& Congenital | Oxx, Pxx, Qxx |
| Psychiatric | Fxx, Gxx |
| Respiratory | Jxx |
| Skin | Lxx |
